# Management and Outcomes of ST-segment Elevation Myocardial Infarction During Coronavirus 2019 Pandemic in a Center with 24/7 Primary Angioplasty Capability: Should We Change Our Practice During Outbreak?

**DOI:** 10.1101/2020.05.02.20088302

**Authors:** Mojtaba Salarifar, Mojgan Ghavami, Hamidreza Poorhosseini, Farzad Masoudkabir, Yaser Jenab, Alireza Amirzadegan, Mohammad Alidoosti, Hassan Aghajani, Ali Bozorgi, Kaveh Hosseini, Masoumeh Lotfi-Tokaldany, Seyedeh Hamideh Mortazavi, Afsaneh Aein, Tahere Ahmadian, Saeed Sadeghian

**Author notes:** **Address correspondence to:** Saeed Sadeghian, MD, Associate Professor of Cardiology, fellowship in Electrophysiology, Cardiovascular Research Institute, Tehran Heart Center, North Kargar-Ave,Tehran-Iran, Postal Code: 1411713138.

## Abstract

**Background:** ST-Elevation Myocardial Infarction (STEMI) is associated with high mortality and morbidity. In order to minimize cardiac tissue injury, primary per-cutaneous coronary intervention (PPCI) as treatment of choice should be performed as soon as possible. Coronavirus Disease 2019 (COVID-19) as an ongoing major global concern affects the other parts of health care system. Applying preventive strategies during this outbreak is necessary. However, critical times in STEMI management and outcomes may be influenced by infection control protocols implementation. The aim of this study is to investigate the differences in time intervals related to STEMI care and 15-day major adverse cardiac events (MACE) during this outbreak compared with the same period in last year and to determine whether the STEMI protocol should be changed to thrombolytic therapy during COVID-19 outbreak or not.

**Methods:** The patients with STEMI who underwent PPCI in Tehran Heart Center were included. Chest Computed tomography (CT) imaging and real time Reverse Transcription Polymerase Chain Reaction (rRT-PCR) were only performed for COVID-19 suspected patients. Seventy-seven patients from 29^th^ February to 29^th^ March 2020 were compared with 62 patients from 1^st^ to 30^th^ March 2019.

**Results:** COVID-19 infection was confirmed by rRT-PCR in 5 cases. CT imaging in 4 out of 5 patients was in favor of COVID-19. The median of door-to-device time was reduced 13 minutes during this outbreak (p: 0.007). In-hospital mortality before and during outbreak was 3.22% and 5.19%, respectively (p: 0.57). Confirmed infection with COVID-19 was only reported in one of expired cases. The difference in 15-day MACE between two time periods was not statistically significant.

**Discussion/Conclusion:** Given that 15-day outcome in acute STEMI patients is not affected by COVID-19 outbreak, we did not find it reasonable to change our protocol. However, further studies are needed to determine a standard protocol for emergency management.

## 1 Introduction

Coronavirus disease 2019 (COVID-19) was first reported in Wuhan, China, on December 2019 but the disease spread rapidly. Presently, almost all the world is affected by COVID-19 ongoing pandemic [1, 2]. COVID-19 has a confirmed human-to-human transmission, even from an asymptomatic carrier, with a basic reproductive number best estimated at 2.2 [2], suggesting that on average, every case of COVID-19 will create 2 new cases. Since the disease is highly contagious and there is no specific treatment for it, prevention through infection control protocols is necessary and inevitable [2–4]. Hospitals and other medical centers are responsible for adapting their inpatient and outpatient healthcare services for this critical condition [5].

Acute ST-segment elevation myocardial infarction (STEMI) is the most acute presentation of coronary artery disease and has considerable morbidity and mortality. In order to minimize the myocardial infarct size and preserve the viability of the ischemic region, it is crucial to expedite the treatment with early diagnosis and quick patient transfer to catheterization laboratory (cath-lab) for primary percutaneous coronary intervention (PPCI) as a definitive treatment [6–8]. However, there is limited data regarding the impact of COVID-19 outbreak on management and outcomes of patients undergoing PPCI [4, 5, 9].

The limited available data shows that COVID-19 outbreak might result in lengthening of the ischemic time in patients with acute STEMI probably due to [4, 5]: 1) implantation of infection control protocol such as utilizing full personal protective equipment (PPE) by PPCI workflow during the treatment process; 2) Asking about the history of travel to contaminated areas, contact with suspected or confirmed cases of COVID-19 and evaluating the symptoms of respiratory infection before transferring the patient to cath-lab and 3) Patients’ disinclination to go to hospital during the COVID-19 outbreak. Given that re-perfusion delay decreases the survival of patients with STEMI, the current pandemic may have negative impact on outcomes of patients [4, 10]. However, to the best of our knowledge, no study has reported the impact of COVID-19 pandemic on outcomes of patients underwent PPCI.

It is well known that PPCI is a more effective revascularization method than thrombolytic therapy in patients with acute STEMI if it can be performed swiftly (within 90 minutes) in capable centers [10]. However, concern regarding the spread of disease from infected patients to PPCI team has resulted in challenging recommendations for choosing reperfusion strategy in acute STEMI patients. To the best of our knowledge, no study has evaluated the efficacy and safety of PPCI during COVID-19 outbreak.

Tehran Heart Center (THC) as a major academic referral hospital for cardiovascular disorders in Iran that provides PPCI service 24 hours a day, 7days a week (24/7) since 2015. On average, 960 PPCI procedures are performed in THC annually. The infection control protocols against COVID-19 have been implemented in this center since 19 February 2020.

In this study we aimed to investigate the impact of COVID-19 outbreak on time components related to STEMI management and 15-day outcomes of patients.

## 2 Materials and Methods

### 2.1 Study participants

All eligible patients who were presented to THC emergency department with acute STEMI from 29^th^ February to 29^th^ March 2020 and underwent PPCI were enrolled in the study. Patients whose diagnosis were not compatible with STEMI by coronary angiography were not included. In addition, the patients who have refused the procedure themselves or the cases in whom cardiac arrest occurred before PPCI (n=0) were excluded. Other exclusion criteria include patients with unclear angina symptoms onset time (n=0) and inpatient candidates (n=2). Ethics committee of

THC and institutional review board approved the study protocol

### 2.2 Protocol

In emergency department, acute STEMI diagnosis is made by the cardiology resident based on electrocardiogram (EKG). Then on-site interventional cardiologist is informed by an immediate telephone call. The cardiology team including cardiology resident and on-site interventional cardiologist then evaluate the patient’s eligibility for PPCI including but not limited to prohibitive comorbidities. If patient is considered eligible for PPCI, the 24/7 code is activated without any further ado and patient is being prepared for transferring to cath-lab after obtaining informed consent. If required, the stat loading dose of aspirin, clopidogrel, statins and other drugs are administered in emergency department. The PPCI procedure is performed if it is indicated according to international guidelines [7, 8].

The STEMI management strategy during COVID-19 outbreak in THC as a referral 24/7 center is displayed in Fig. 1. All patients are evaluated regarding sign and symptoms of infection with COVID-19 using a pre-formed checklist (Table 1). For COVID-19 suspected patients, spiral chest computed tomography (CT) is performed after PPCI and nasal and oropharyngeal swab samples are also collected for nucleic acid testing by real time Reverse Transcription Polymerase Chain Reaction (rRT-PCR) utilizing Nanjing Vazyme Medical Technology Co., Ltd (2019-nCoV) Triplex RT-qPCR Detection Kit (CE-IVD). Unfortunately, due to shortage of RT-PCR kits ‘’test-all’’ strategy was not performed for all admitted patients.

**Fig. 1.**
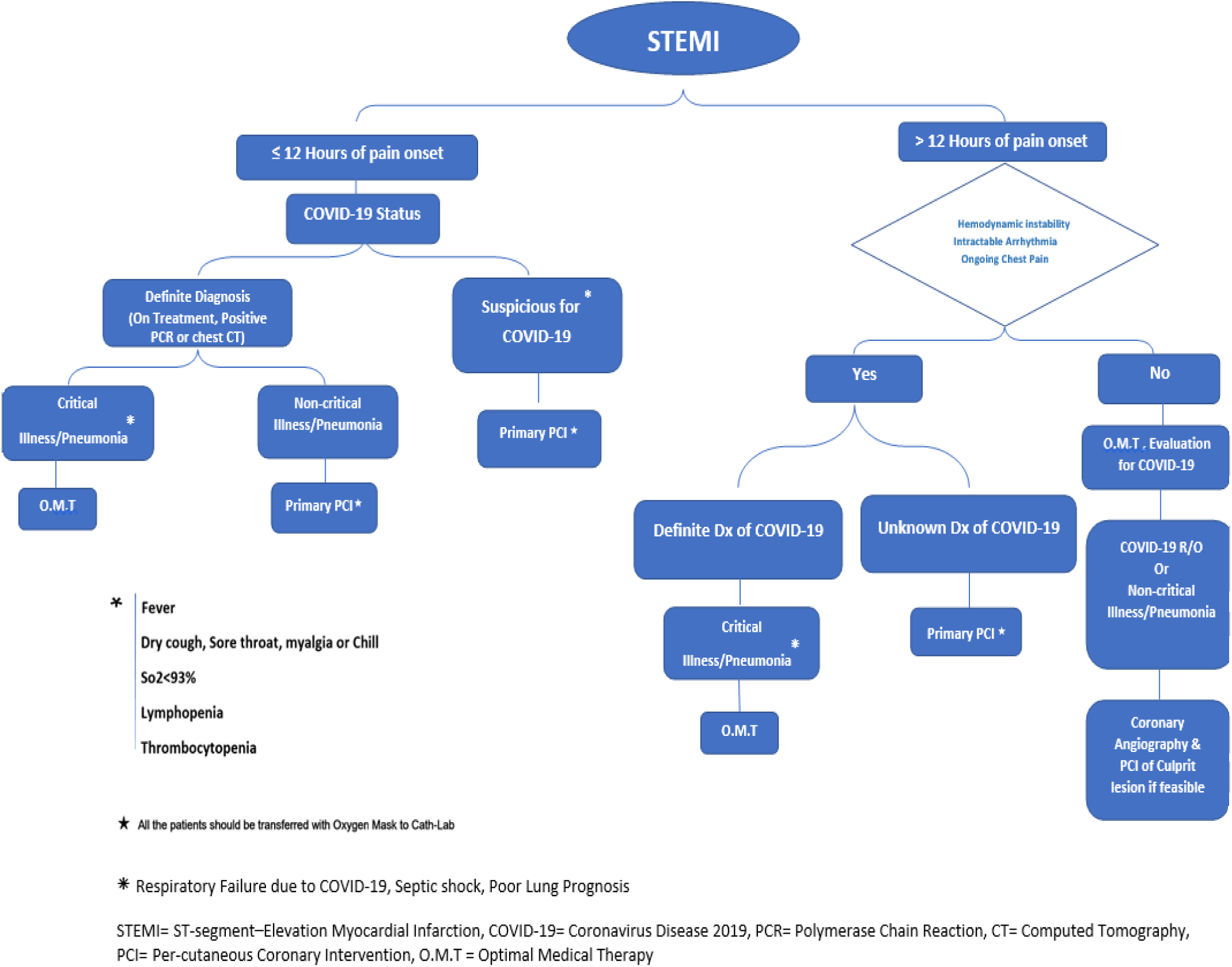
Tehran Heart Center ST-Elevation Myocardial Infarction management protocol during Coronavirus Disease 2019 outbreak

**Table 1.** Screening corona virus disease 2019 check list

|  |  |
| --- | --- |
| Fever ($>37.3^{\circ}\text{C}$) | Head-ache |
| History of direct contact with COVID-19 +<br>within the last 2 weeks | Diarrhea |
| Sore throat | Chill |
| Dry cough | Dyspnea |
| Myalgia | Anosmia |
| Rhinorrhea or sneezing | Ageusia |

Given that the patients may be asymptomatic carriers of COVID-19, all the cath-lab staff in close contact with the patient wear appropriate PPE regardless of respiratory infection signs and symptoms. PPE include an isolated gown, disposable gloves, a face shield or goggles and a N95 mask for each procedure.

All health care personnel are evaluated daily and are asked about their symptoms. Body temperature was checked routinely.

To determine the impact of COVID-19 outbreak on STEMI care related time components and 1598 day outcome in these cases, we compared them to the patients with acute STEMI who were treated by PPCI from 1st March to 30th March 2019 (n=62) using THC 24/7 data bank registry. The same inclusion and exclusion criteria were considered for these patients. To follow up the patients who were discharged (from 1 to 30 days previously) during this outbreak, trained nurses made a telephone call to all of them and recorded their current symptoms, living status and major adverse cardiac event (MACE) components.

### 2.3 Definitions

We evaluated time components related to STEMI care including symptom onset-to-door, door-to-device, symptom onset-to-device. The time when the patient arrives at the emergency department and the time of successful guidewire passage during PPCI are defined as door and device time, respectively.

A 15-day MACE was evaluated as the outcome of STEMI patients and was defined as all-cause mortality, non-fatal MI, repeated revascularization (PCI or coronary artery bypass grafting), cerebrovascular accidents and hospitalization.

### 2.4 Satistical analysis

We reported categorical variables as percentages and continuous variables as means (±standard deviations (SD)) and medians (IQR25-75%) for those with and without normal distribution, respectively. To assess normality distribution of variables Kolmogorov-Smirnov test was conducted.

Group differences analyses were performed using independent t-test, Mann–Whitney U test and chi-square test for continuous (with/ without normal distribution) and categorical variables, respectively.

The statistical data analysis was performed using software SPSS, version 26 and P value < 0.05 was defined as significant.

## 3 Results

Between 29^th^ February and 29^th^ March 2020, 77 patients with STEMI who underwent PPCI, were compared with 62 STEMI patients during the same time period in the previous year. Table 2 shows demographic and clinical characteristics of the patients. There were no significant differences between the two groups except for borderline significant higher prevalence for male sex among patients treated during COVID-19 outbreak (86.7% vs 72.6%, p = 0.05). Chest CT scan and rRT-PCR nucleic acid testing were performed for 15 patients who were considered suspected based on checklist. From 8 cases with chest CT imaging in favor of COVID-19, the infection was confirmed by rRT-PCR testing in 4 patients. Serial chest CT scans in one patient with suspicious symptoms revealed normal findings, while rRT-PCR testing confirmed the infection in this patient.

**Table 2.**
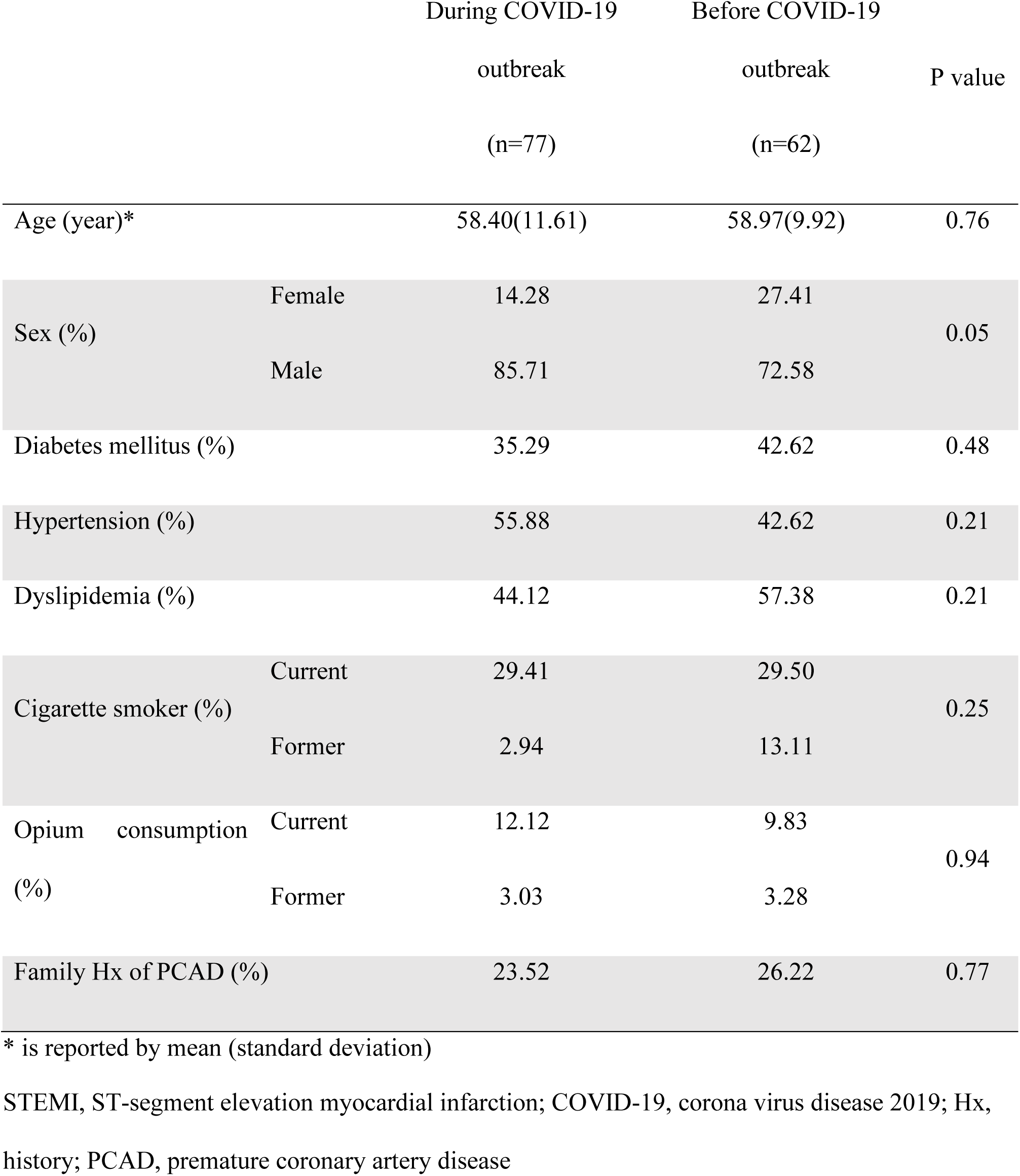
Demographic and clinical characteristics of the patients with acute STEMI that underwent primary coronary angioplasty during COVID-19 outbreak (29 ^th^ February to 29 ^th^ March 2020) and before COVID-19 outbreak (1^st^ to 30^th^ March 2019)

As shown in Table 3, door-to-device time was shorter during COVID-19 outbreak compared to the previous year (47.0 vs 60.0 min, p = 0.00). No significant differences were observed between the two groups with respect to symptom-to-door and symptom-to-device times.

**Table 3.**
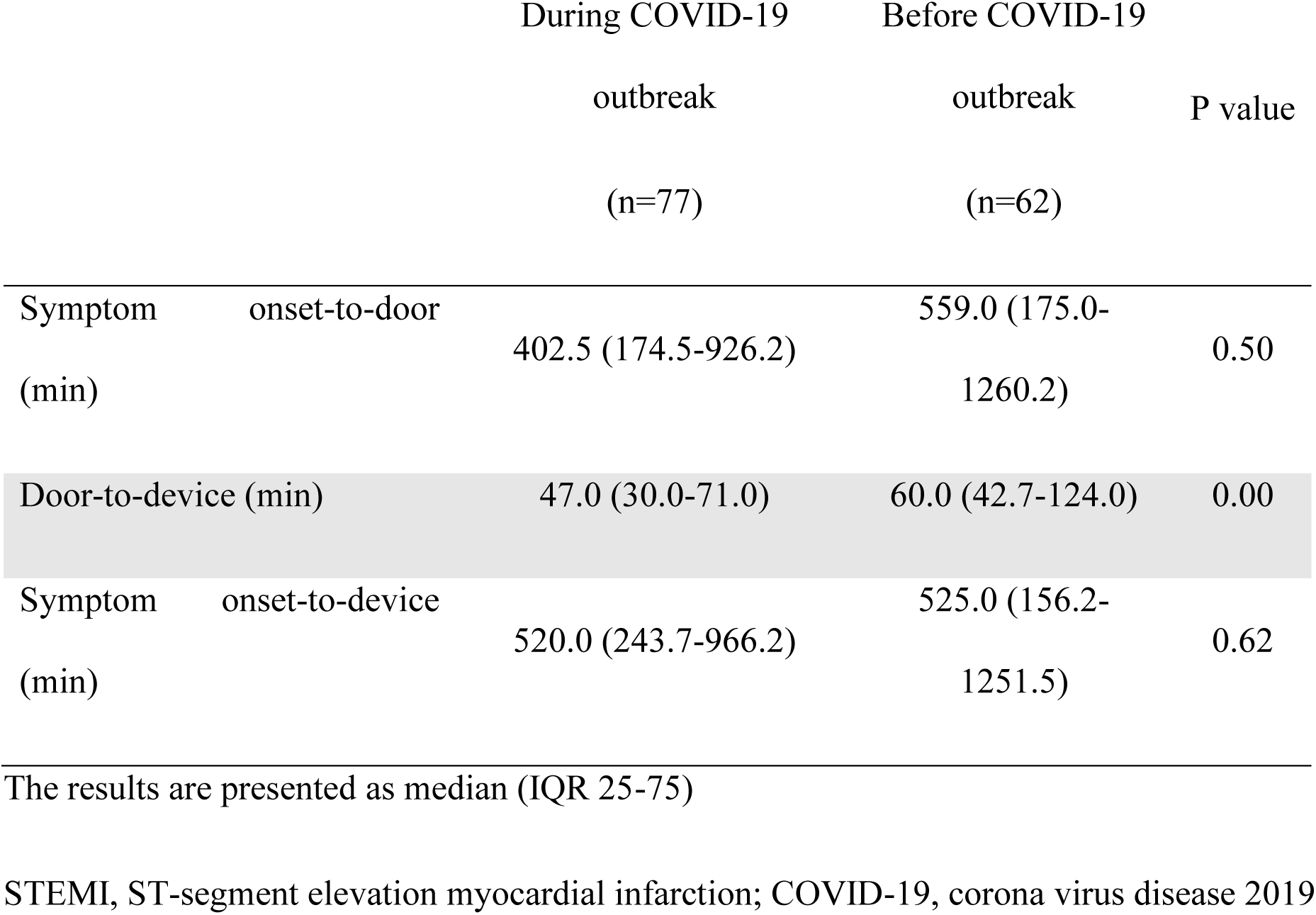
Critical time intervals for reperfusion in patients with acute STEMI during COVID-19 outbreak (29^th^ February to 29^th^ March 2020) and before COVID-19 outbreak (1^st^ to 30^th^ March 2019)

There was no significant difference between patients treated during COVID-19 outbreak (4 patients, 5.2%) and those treated during the same period in previous year (2 patients, 3.2%) with respect to 15-day all-cause mortality (p = 0.57). While last year 2 patients developed MACE (1 non-fatal MI, 1 repeated revascularization with coronary artery bypass graft (CABG)), none of the patients had non-fatal MACE during the COVID-19 outbreak. There was no statistically significant difference between the two groups with respect to 15-day MACE (Table 4). None of the cath lab personnel presented with COVID-19 related symptoms until the end of the study.

**Table 4.**
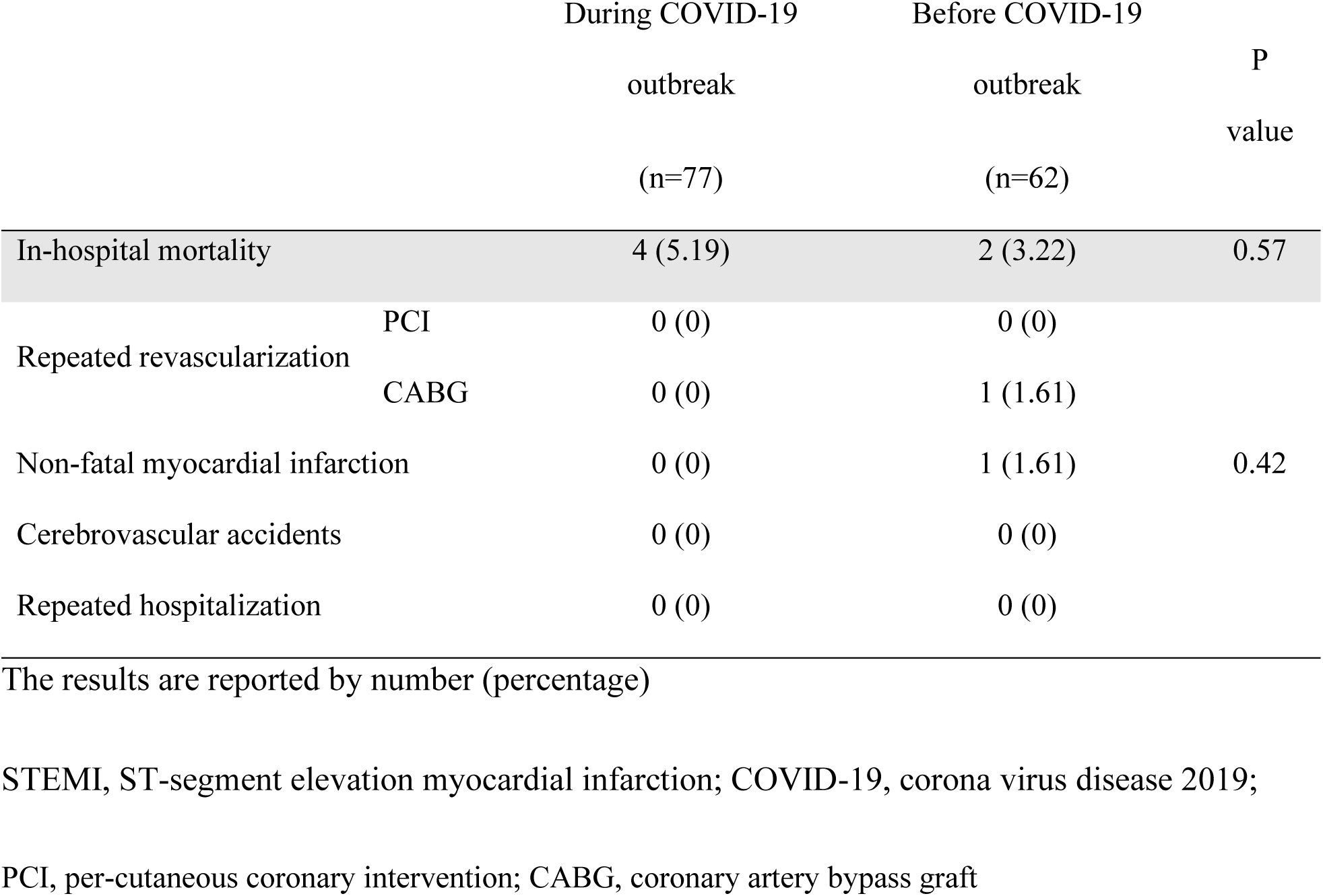
Fifteen-day major adverse cardiac events in patients with STEMI underwent primary coronary angioplasty before and during COVID-19 outbreak

One out of 5 patients with rRT-PCR-confirmed COVID-19 expired during hospitalization. The case was a 48-year-old male with the history of diabetes mellitus, hypertension and kidney transplantation 9 years ago, and was receiving immunosuppressive drugs. He was also cigarette smoker and opium abuser. The patient presented with sore throat and weakness in addition to typical chest pain. The symptom-onset to door and door-to-device time intervals were 149 minutes and 30 minutes, respectively. His coronary angiography demonstrated left anterior descending (LAD) cut-off at mid part, and 60% stenosis in proximal part of right coronary artery (RCA) and obtuse marginal (OM). Successful PPCI was done with providing full PPE for cath-lab personnel. COVID-19 infection was confirmed with spiral chest CT imaging (shown in Fig. 2) and the rRT-PCR nucleic acid testing on day of admission. Supportive care including ventilation and routine empiric drugs (Hydroxychloroquine and Lopinavir/Ritonavir) was started. The day after procedure, the patient complained of dyspnea. Echocardiography was done immediately; ejection fraction was 40% with no significant valvular heart disease and no pericardial effusion. His oxygen saturation decreased to 93%. However, his condition deteriorated rapidly; ventricular fibrillation occurred. Unfortunately, cardiopulmonary resuscitation was unsuccessful. Long “QT interval” was not detected during patients’ hospitalization. _Creatinine level was 3 mg/dL and other electrolytes were normal. Severe hypoxia and acute respiratory disease in the context of COVID-19 may be the cause of his death.

**Fig. 2.**
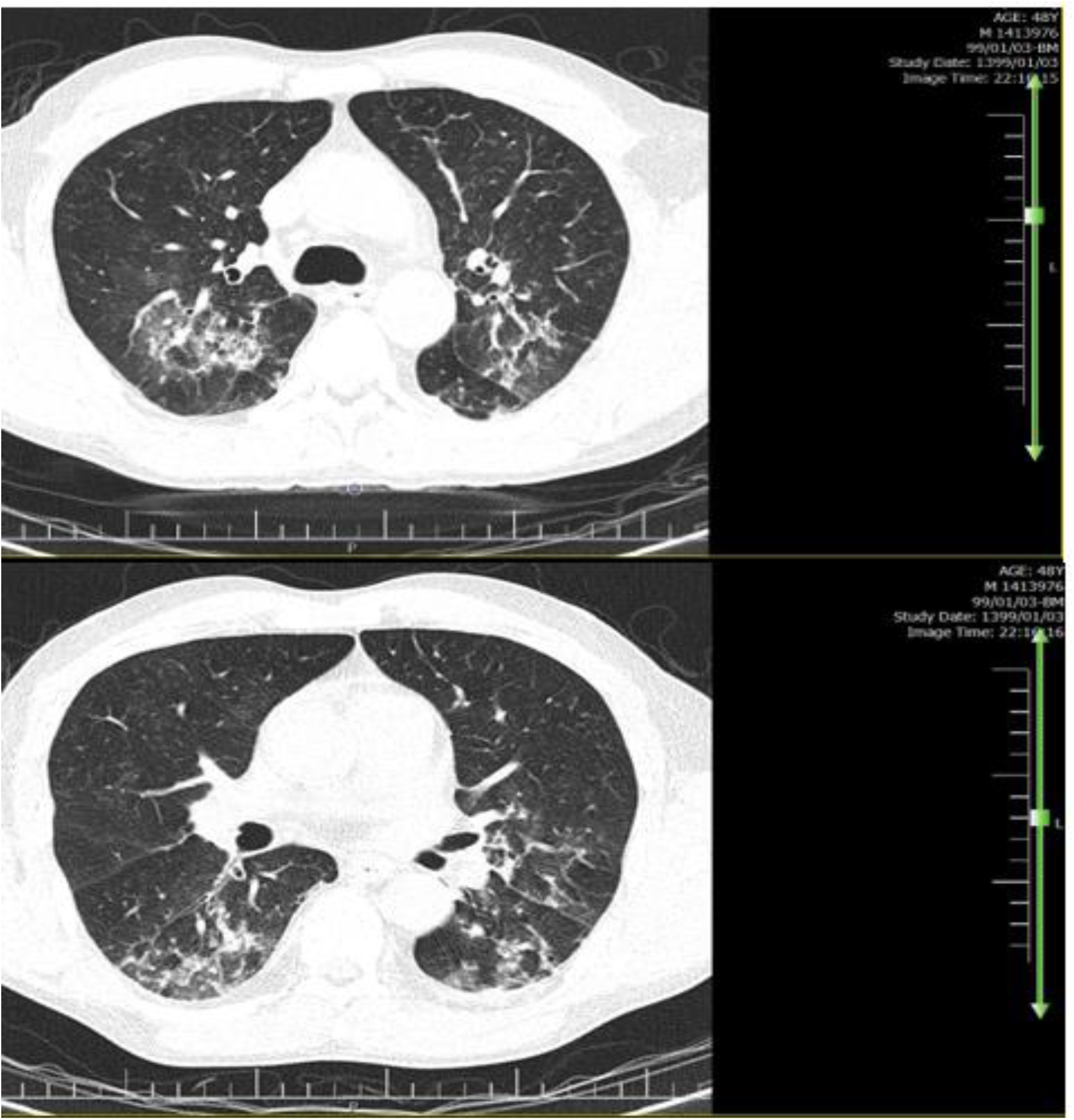
Spiral chest computed tomography findings in the expired patient with confirmed coronavirus disease 2019

## 4 Discussion/ Conclusion

Our study demonstrated that during COVID-19 outbreak the critical times for reperfusion in STEMI patients undergoing PPCI not only did not increase but also the door-to-device time reduced significantly. Moreover, outcomes of patients in terms of 15-day mortality and 15-day MACE did not differ significantly during the COVID-19 outbreak compared to same period in 2019.

Considering the vast dimensions of COVID-19, the impact of the disease on emergency management such as acute MI is largely obscure.

In this study we observed a trend toward reduced time intervals in all critical times for reperfusion in STEMI patients undergoing PPCI during COVID-19 outbreak and a statistically significant reduction was observed with respect to door-to-device time. This finding contrasts to a recent study by Tam et al from Hong Kong [4]. They compared STEMI related time components between 7 patients with STEMI who underwent PPCI within 15 days during COVID-19 outbreak and 108 patients with similar condition during the previous year and observed an increase in all the time intervals during outbreak especially symptom onset-to-first medical contact. Tam and colleagues believe that these differences are related to patients’ unwillingness to go to hospital during COVID-19 outbreak and the time spent for infection protocol implementation [4]. However, in our study we did not observe a prolongation in symptom onset-to-door time. This finding may be justified by a delay because of unwillingness to go to hospital, which is balanced with early arrival to hospital due to significantly unloaded traffics of city during outbreak. Meanwhile, it seems that limited elective invasive procedures during COVID-19 outbreak has resulted in reduced waiting time for preparation of routinely occupied cath-labs and diminishing the cath-lab staff workload with subsequent performance improvement that led to door-to-device time interval shortening. This was observed, even though infection control protocol and PPE were routinely used for all patients.

Concerns regarding spread of the disease have resulted in some recommendations and adaptations in routine management of STEMI patients. A Chinese protocol [11] suggests fast nucleic acid testing to detect suspected or confirmed infected cases prior to deciding about reperfusion strategy. They suggest thrombolytic therapy for suspected/confirmed COVID-19 cases with mild/moderate pneumonia who present to emergency department within 12 hours from symptom onset. However, this approach has been challenged in communities such as the United States in which PPCI is a routine revascularization method. Additionally, rapid nucleic acid testing for all patients is not easily available. American College of Cardiology’s (ACC) Interventional Council and the Society of Cardiovascular Angiography and Intervention (SCAI) have recommended fibrinolytic therapy in relatively stable patients with STEMI and active confirmed COVID-19 as an option. They recommend appropriate PPE if PPCI is to be performed in those cases [5].

In this study we demonstrated that outcomes of patients undergoing PPCI during COVID-19 outbreak is the same as the past. Meanwhile, using PPE equipment, none of the cath-lab personnel was infected despite performing PPCI for 8 patients with suggestive chest CT scans for COVID-19 and 5 patients with confirmed COVID-19 infections using rRT-PCR test.

Taking together, if full PPE can be provided for PPCI team, using the protocol of THC for management of patients with STEMI (shown in Fig. 1) might be a more efficient, practical and evidence-based approach compared to recommendations for using thrombolytic therapy in patients with suspected/confirmed COVID-19 infection. However, further studies are needed to determine a standard protocol for management of patients with acute STEMI during COVID-19 outbreak.

## Data Availability

All the data are available on an open repository.

## 5 Acknowledgment

None declared.

## 6 Statement of Ethics

The research was conducted ethically in accordance with the World Medical Association Declaration of Helsinki. Subjects have given their written informed consent. The study protocol was approved by the institute’s committee on human research (IR.TUMS.VCR.REC.1399.023).

## 9 Author Contributions

“Mojgan Ghavami” conceived the original idea, performed data entry and analysis and wrote the manuscript with support from “Farzad Masoudkabir”, “Kaveh Hosseini” and “Seyedeh Hamideh Mortazavi”. “Mojtaba Salarifar” and “Saeed Sadeghian” supervised the project. “Hamidreza Poorhosseini”, “Yaser Jenab”, “Alireza Amirzadegan”, “Mohammad Alidoosti”, “Hassan Aghajani”, “Ali Bozorgi” and “Masoumeh Lotfi-Tokaldany” helped supervise the project. “Afsaneh Aein” and “Tahere Ahmadian” conducted the data collection and data entry. All authors discussed the results and contributed to the final manuscript.

